# TEMPORAL TRENDS AND TRANSMISSION DYNAMICS OF PRE-TREATMENT HIV-1 DRUG RESISTANCE WITHIN AND BETWEEN RISK GROUPS IN KENYA, 1986-2020

**DOI:** 10.1101/2023.03.21.23287487

**Authors:** George M. Nduva, Frederick Otieno, Joshua Kimani, Yiakon Sein, Dawit A. Arimide, Lyle R. McKinnon, Francois Cholette, Morris K. Lawrence, Maxwell Majiwa, Moses Masika, Gaudensia Mutua, Omu Anzala, Susan M. Graham, Larry Gelmon, Matt A. Price, Adrian D. Smith, Robert C. Bailey, Patrik Medstrand, Eduard J. Sanders, Joakim Esbjörnsson, Amin S. Hassan

## Abstract

**Background:** Evidence on the distribution of pre-treatment HIV-1 drug resistance (HIVDR) by risk groups is limited in Africa. We assessed prevalence, trends, and transmission dynamics of pre-treatment HIVDR within-and-between men who have sex with men (MSM), people who inject drugs (PWID), female sex workers (FSW), heterosexuals (HET), and children infected perinatally in Kenya.

**Methods:** HIV-1 partial *pol* sequences from antiretroviral-naïve samples collected between 1986-2020 were used. Pre-treatment RTI, PI and INSTI mutations were assessed using the Stanford HIVDR database. Phylogenetics methods were used to determine and date transmission clusters.

**Results:** Of 3567 sequences analysed, 550 (15.4%, 95% CI: 14.2-16.6) had at least one pre-treatment HIVDR mutation, which was most prevalent amongst children (41.3%), followed by PWID (31.0%), MSM (19.9%), FSW (15.1%) and HET (13.9%). No INSTI resistance mutations were detected. Among HET, pre-treatment HIVDR increased from 6.6% in 1986-2005 to 20.2% in 2011-2015 but dropped to 6.5% in 2016-2020. Overall, 22 clusters with shared pre-treatment HIVDR mutations were identified. The largest was a K103N mutation cluster involving 16 MSM sequences sampled between 2010-2017, with an estimated tMRCA of 2005 (HPD, 2000-2008). This lineage had a growth rate=0.1/year and R_0_=1.1, indicating propagation over 12 years among ART-naïve MSM in Kenya.

**Conclusions:** Compared to HET, children and key populations had higher levels of pre-treatment HIVDR. Introduction of INSTIs after 2016 may have reversed the increase in pre-treatment RTI mutations in Kenya. Continued surveillance of HIVDR, with a particular focus on children and key populations, is warranted to inform treatment strategies in Kenya.

**Summary:** Compared to the heterosexual population, key populations had higher levels of pre-treatment HIV-1 drug resistance (HIVDR). Propagation of HIVDR was risk-group exclusive. Introduction of integrase inhibitors abrogated propagation of reverse transcriptase inhibitors mutations among the heterosexual, but not key populations.

## INTRODUCTION

By the end of 2020, an estimated 30 million individuals were receiving antiretroviral therapy (ART) globally^1^. The scale-up of ART has substantially reduced new HIV-1 infections, vertical transmissions, HIV-1 related mortality, and improved life expectancy for millions of people with HIV (PWHIV)^1-3^. However, widespread use of ART at the population-level has been associated with the emergence of HIV-1 drug resistance (HIVDR), and the transmission of drug resistant viruses that can compromise therapy outcomes^4-9^.

The World Health Organization (WHO) recommends routine nationally representative HIVDR surveys to inform treatment strategies^10,11^. Increasing levels of pre-treatment HIVDR to non-nucleoside reverse transcriptase inhibitors (NNRTIs) has been recognized as a problem for many years, especially in low and middle-income countries (LMIC) where virologic monitoring of patients on ART is sub-optimal^12-15^. Out of concern for treatment failure in the setting of increasing NNRTI resistance and poor tolerability of PIs, WHO has prompted a switch to INSTIs with a high genetic barrier as first-line regimens in LMIC settings – and Dolutegravir (DTG) is now widely adopted in first-line regimen both for treatment naïve and experienced PWHIV^16,17^. However, it is not clear if the switch to DTG-based regimens, which came into effect in LMIC settings in 2016, may have impacted emergence and circulation of NNRTI resistant strains and this warrants further evaluation.

Further, key populations including men who have sex with men (MSM), people who inject drugs (PWID), female sex workers (FSW), transgender people, and people in prisons, are disproportionately impacted by HIV-1 in Sub-Saharan Africa. Transmission from key population individuals is thought to contribute disproportionately to Africa’s generalized HIV-1 epidemic^18,19^. We previously demonstrated that the majority of HIV-1 transmissions in Kenya occur within distinct risk groups,^20-22^ and, although infrequent, HIV-1 transmission from HET to key populations is more common than vice-versa^22^. However, it remains unknown whether there is differential transmission of HIVDR mutations within and between HET and key populations in Kenya. This study aimed to describe prevalence, temporal trends, and transmission linkages of pre-treatment HIVDR mutations within and between risk groups in Kenya.

## METHODS

### Study population

HIV-1 partial *pol* sequences from Kenya were either newly generated from archived plasma samples (henceforth referred as newly generated) or retrieved from the Los Alamos HIV-1 sequence database (henceforth referred as published sequences)^23^. The newly generated HIV-1 *pol* sequences (approximately 1020 nucleotides, HXB2 [K03455] positions 2267-3287) were processed from blood plasma as previously described^24^. These samples were obtained through the MSM Health Research Consortium – a multi-site collaboration between the KEMRI-Wellcome Trust Research programme (KWTRP) in Coastal Kenya^25,26^, Nyanza Reproductive Health Society (NRHS) in Western Kenya^27^, Sex Workers Outreach Program (SWOP) clinics in Nairobi, Kenya and the TRANSFORM MSM cohort from Nairobi, Kenya^28^, and from the national HIV-1 reference laboratory at the KEMRI-CGHR in Kisumu, Kenya. Samples identified between 2006-2011 were collected under IAVI’s Protocol C, a study of incident HIV infection among higher risk persons; this included teams in Nairobi and KWRTP^29^. The published Kenyan HIV-1 *pol* sequences (corresponding to the HXB2 - K03455, *protease* and *reverse transcriptase* nucleotide positions 2000-4800 and *integrase* positions 4230-5093) were retrieved from the Los Alamos HIV database on March 20^th^ 2022^23^.

Newly generated and published sequences were annotated with sampling date, sampling location (county and province), ART status (whether treatment naïve or experienced), age, sex, and risk group (MSM; PWID; FSW; children, and HET). Where demographic data were missing for published sequences, information was obtained from reviewing associated literature or through direct communication with study authors. Sequences from ART-experienced individuals were excluded from this analysis.

### HIV-1 subtype determination

Newly generated and published partial *pol* sequences from Kenya were aligned with the HIV-1 Group M (subtypes A-K + Recombinants) subtype reference sequences (http://www.hiv.lanl.gov) using the MAFFT algorithm in Geneious Prime 2019^23^. Subtypes were determined by maximum-likelihood (ML) phylogenetic analysis in PhyML using the general time-reversible substitution model with a gamma-distributed rate variation and proportion of invariant sites (GTR+Γ4+I)^30^. Branch support was determined using the approximate likelihood ratio test with the Shimodaira-Hasegawa-like procedure (aLRT-SH) in PhyML and aLRT-SH ≥0.90 was considered significant. The phylogeny was visualized using FigTree v1.4.4 (https://github.com/rambaut/figtree/releases). Unique recombinant forms (URFs) were characterised by boot-scan analysis in SimPlot^31-33^. Subtype assignment was also compared with results from the REGA subtyping tool (version 3.0).

### Characterisation of pre-treatment HIV-1 drug resistance

All sequences from treatment-naïve individuals were submitted to the Stanford HIV database (HIVdb Algorithm, version 8.8) for determination and interpretation of HIVDR mutations. The Calibrated Population Resistance tool was used to identify pre-treatment drug resistance mutations, and the results cross-referenced with the WHO list of mutations for the surveillance of transmitted HIVDR^34,35^. Mutations were grouped and presented based on drug class as follows: nucleoside reverse transcriptase inhibitors (NRTI), non-nucleoside reverse transcriptase inhibitors (NNRTI), protease inhibitors (PI) and integrase strand transfer inhibitors (INSTI) mutations.

### Phylogenetic determination of clusters and Bayesian inference

HIV-1 sequences from ART-naïve individuals were grouped into the main HIV-1 subtypes observed (A1, C and D). For each subtype-specific dataset, a search for related sequences was done separately using the NCBI GenBank BLAST tool, with results limited to a threshold of 10 similar hits per individual-specific sequence, as previously described^21,36,37^. Reference sequences and sequences in the Kenyan dataset with pre-treatment HIVDR mutations were compiled into subtype-specific alignments, excluding codon positions associated with drug resistance mutations. ML phylogenies were generated in PhyML using the GTR+Γ4+I model^30^ and phylogenetic trees were annotated based on the most dominant mutations per drug class i.e. K103N/S, Y181C/I/V, and G190A/S/E for NNRTI resistance and M184V/I and T215 revs for NRTI resistance. Kenyan HIV-1 clusters were defined by aLRT-SH support ≥0.9 and having ≥80% Kenyan sequences, and then further classified by number of sequences into dyads (2 sequences), networks (3-14) sequences, and large clusters (>14 sequences)^21,36,37^.

Dating of clusters were performed in BEAST (version 1.10.4) using the GTR+Γ4+I substitution model, an uncorrelated lognormal relaxed clock under a Skygrid coalescent tree prior, and specifying a hierarchical phylogenetic model (HPM) to enhance precision in estimating evolutionary parameters^38,39^. Cluster growth rates were inferred under a logistic growth model^40-42^. BEAST runs of 300 million generations were computed, sampling every 30,000^th^ iteration, and discarding the first 10% as chain burn-in. Convergence was determined in Tracer v.1.7.0 (defined as effective sample sizes (ESS) ≥100)^38^. Maximum clade credibility (MCC) trees were summarized from the posterior tree distribution using Tree-Annotator v1.10.4 (BEAST suite) and visualized using Figtree (v1.4.4, available at: https://github.com/rambaut/figtree/releases). The basic reproductive number (R_0_, defined as the number of secondary infections that arise from a typical primary case in a completely susceptible population) per cluster was estimated based on the respective cluster growth rate (*r*) using the formula R_0_□=□*r*D+1 (where D represents the average duration of infectiousness for individuals) ^40-42^. In the absence of empiric data on the duration of infectiousness in our setting, we assumed D to be one year.

### Statistical analysis

Sequences were categorized by year of sampling and calendar period collapsed into five-year intervals to reflect major changes in treatment guidelines in Kenya as follows: before 2005 (before introduction of combination ART [cART]), 2005-2010 (introduction of cART), 2011-2015 (scale up of cART), and 2016-2020 (introduction of INSTI-based regimen). Categorical data were compared with the χ^2^ test and continuous data with the Kruskal-Wallis test where appropriate. Overall, drug-class-specific, and mutation-specific HIVDR prevalence estimates and 95% confidence interval (CI) were presented. HIVDR temporal trends were assessed using *nptrends*, a nonparametric extension of the Wilcoxon rank-sum test^43^. Multivariable logistic regression models were used to determine associations between risk groups, geographic locations, sampling period and pre-treatment HIVDR. All statistical tests were 2-tailed (P<0.05). Data analysis and summary plots were done using Stata 15 (StataCorp LLC, College Station, Texas, USA) and RStudio (version 1.2.5001) with the *ggplot2* package^44^.

### Nucleotide sequence accession numbers

Newly generated nucleotide sequences are available from GenBank under the accession numbers OM109695-OM110282.

### Ethical consideration

For the newly generated sequences, informed consent for use of data and samples for research studies was obtained from participants under respective study protocols. Since published sequences were obtained from an open access repository, informed consent was not retrospectively obtained. Instead, science and ethics approval were obtained from the Kenya Medical Research Institute (KEMRI) Scientific and Ethics Review Unit (SERU 3547). All newly generated and published sequences were de-identified at source before inclusion in the current analysis.

## RESULTS

### Study population

Overall, 5572 HIV-1 *pol* sequences (n=755 newly generated, and n=4817 published) sampled between 1986-2020 were considered. Sequences from ART-experienced individuals (n=1778) and those with unverified treatment status (n=227) were excluded from the analysis (Figure 1). The analysed dataset comprised 3567 HIV-1 sequences from ART-naïve individuals covering the reverse transcriptase (n=3567, 100.0%), protease (n=2491, 69.8%) and integrase (n=106, 3.0%) coding regions. Most sequences were from the HET population (n=2947, 82.1%) and from the former Nairobi (n=1479, 41.5%) and Coast (n=1027, 28.8%) provinces (Table 1). The most common circulating strain was HIV-1 sub-subtype A1 (n=2282, 64.0%), followed by subtype D (n=509, 14.3%), unique recombinant forms (URFs, n=413, 11.6%), and subtype C (n=282, 7.9%), with this distribution being mostly similar across risk groups (Table 2).

**Table 1.** Distribution of newly generated and published HIV-1 *pol* sequences from antiretroviral therapy-naïve individuals in Kenya (n=3567, 1986-2020).

| Characteristics |  | Newly generated<br>n=396 (%) | Published<br>N=3,171 (%) | Total<br>N=3,567 (%) |
| --- | --- | --- | --- | --- |
| Risk group | HET | 78 (19.7%) | 2869 (90.5%) | 2947 (82.6%) |
|  | MSM | 190 (48.0%) | 151 (4.8%) | 341 (9.6%) |
|  | PWID | 0 (0.0%) | 58 (1.8%) | 58 (1.6%) |
|  | FSW | 128 (32.3%) | 18 (0.6%) | 146 (4.1%) |
|  | Children | 0 (0.0%) | 75 (2.4%) | 75 (2.1%) |
| Year (range) | Before 2005 | 18 (4.5%) | 433 (13.7%) | 451 (12.6%) |
|  | 2006-2010 | 110 (27.8%) | 1887 (59.5%) | 1997 (56.0%) |
|  | 2011-2015 | 41 (10.4%) | 842 (26.6%) | 883 (24.8%) |
|  | 2016-2020 | 227 (57.3%) | 9 (0.3%) | 236 (6.6%) |
| Province | Nairobi | 150 (37.9%) | 1329 (41.9%) | 1479 (41.5%) |
|  | Coast | 175 (44.2%) | 852 (26.9%) | 1027 (28.8%) |
|  | Nyanza | 71 (17.9%) | 662 (20.9%) | 733 (20.5%) |
|  | Rift Valley | 0 (0.0%) | 294 (9.3%) | 294 (8.2%) |
|  | Central | 0 (0.0%) | 33 (1.0%) | 33 (0.9%) |
|  | North-Eastern | 0 (0.0%) | 1 (0.0%) | 1 (0.0%) |

*Abbreviations: ART, combined antiretroviral therapy; HET, presumed heterosexual i.e., men and women not reporting sex work or male same-sex behaviour; MSM, men who have sex with men; PWID, people who inject drugs; FSW, female sex worker.*

**Table 2.**
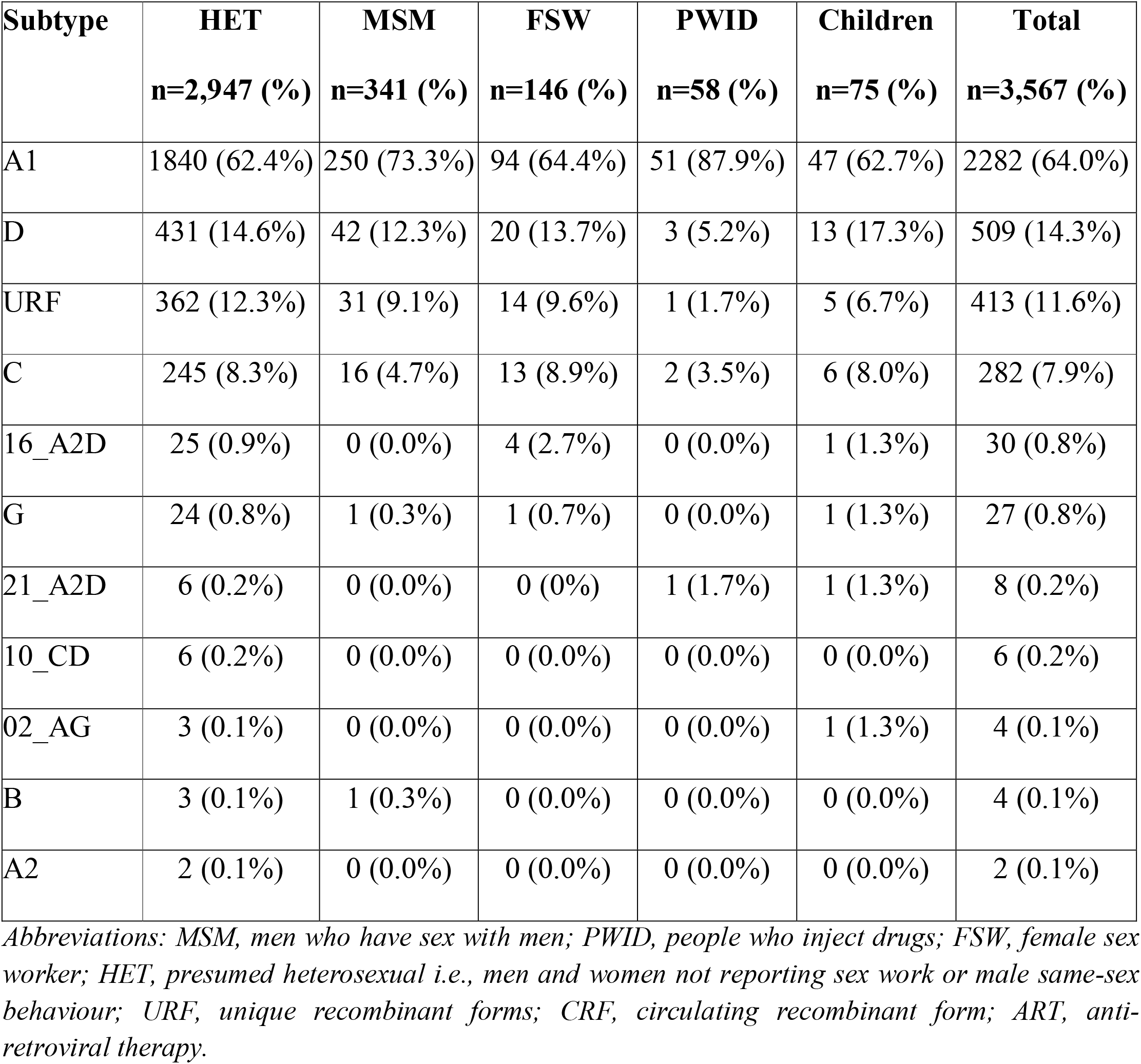
HIV-1 subtypes distribution within-and-between risk groups in Kenya (n=3567, 1986-2020).

| Subtype | HET<br>n=2,947 (%) | MSM<br>n=341 (%) | FSW<br>n=146 (%) | PWID<br>n=58 (%) | Children<br>n=75 (%) | Total<br>n=3,567 (%) |
| --- | --- | --- | --- | --- | --- | --- |
| A1 | 1840 (62.4%) | 250 (73.3%) | 94 (64.4%) | 51 (87.9%) | 47 (62.7%) | 2282 (64.0%) |
| D | 431 (14.6%) | 42 (12.3%) | 20 (13.7%) | 3 (5.2%) | 13 (17.3%) | 509 (14.3%) |
| URF | 362 (12.3%) | 31 (9.1%) | 14 (9.6%) | 1 (1.7%) | 5 (6.7%) | 413 (11.6%) |
| C | 245 (8.3%) | 16 (4.7%) | 13 (8.9%) | 2 (3.5%) | 6 (8.0%) | 282 (7.9%) |
| 16_A2D | 25 (0.9%) | 0 (0.0%) | 4 (2.7%) | 0 (0.0%) | 1 (1.3%) | 30 (0.8%) |
| G | 24 (0.8%) | 1 (0.3%) | 1 (0.7%) | 0 (0.0%) | 1 (1.3%) | 27 (0.8%) |
| 21_A2D | 6 (0.2%) | 0 (0.0%) | 0 (0%) | 1 (1.7%) | 1 (1.3%) | 8 (0.2%) |
| 10_CD | 6 (0.2%) | 0 (0.0%) | 0 (0.0%) | 0 (0.0%) | 0 (0.0%) | 6 (0.2%) |
| 02_AG | 3 (0.1%) | 0 (0.0%) | 0 (0.0%) | 0 (0.0%) | 1 (1.3%) | 4 (0.1%) |
| B | 3 (0.1%) | 1 (0.3%) | 0 (0.0%) | 0 (0.0%) | 0 (0.0%) | 4 (0.1%) |
| A2 | 2 (0.1%) | 0 (0.0%) | 0 (0.0%) | 0 (0.0%) | 0 (0.0%) | 2 (0.1%) |

*Abbreviations: MSM, men who have sex with men; PWID, people who inject drugs; FSW, female sex worker; HET, presumed heterosexual i.e., men and women not reporting sex work or male same-sex behaviour; URF, unique recombinant forms; CRF, circulating recombinant form; ART, anti-retroviral therapy.*

**Figure 1.**
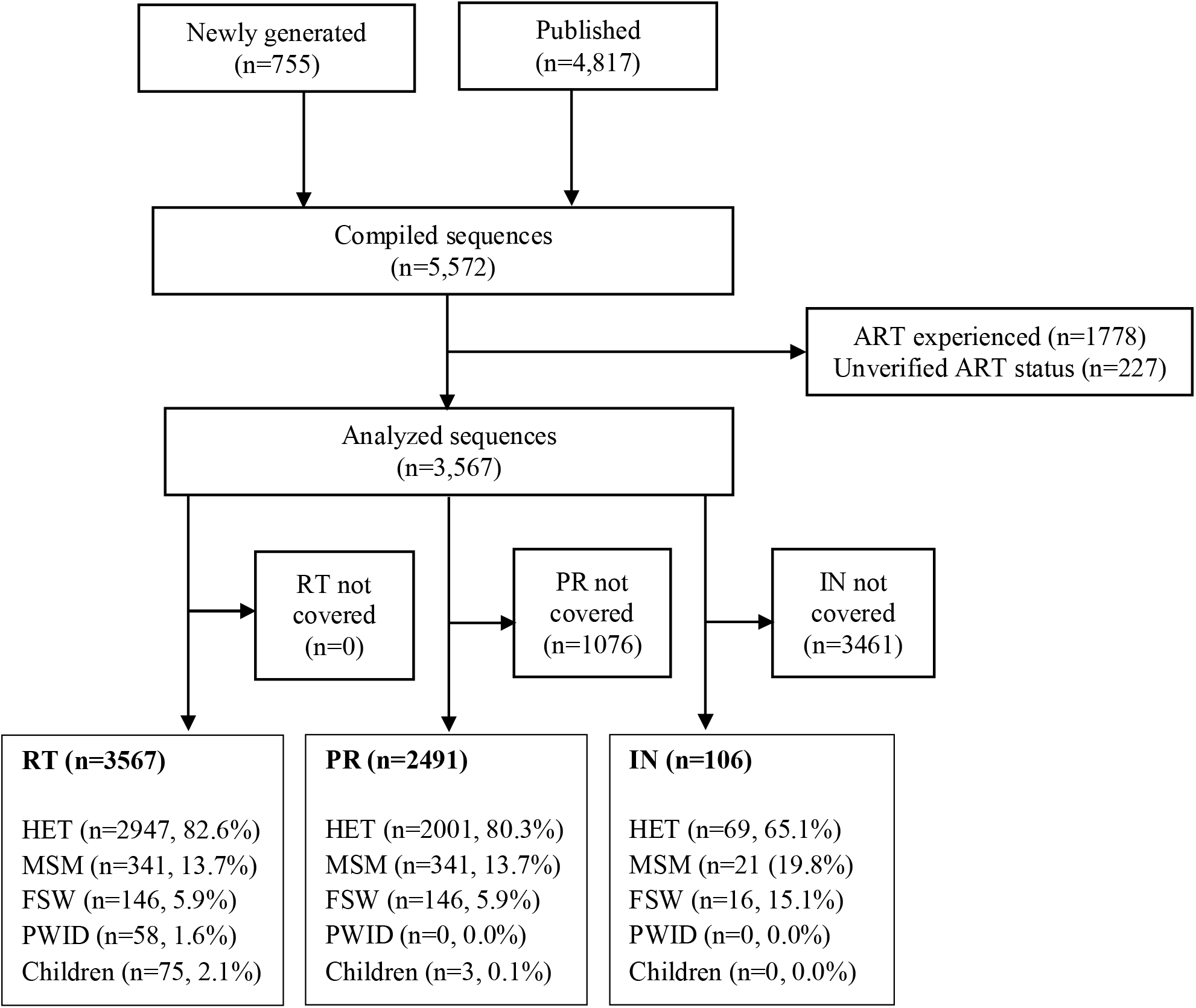
A flowchart summarising the number of HIV-1 pol sequences (1986-2020) analysed in this study. Abbreviations: ART, antiretroviral therapy; RTI, reverse-transcriptase inhibitors; PI, protease inhibitors; INSTI integrase inhibitors; MSM, men who have sex with men; PWID, people who inject drugs; FSW, female sex worker; HET, at-risk men and women who did not report sex work or male same-sex behaviour.

### Prevalence of pre-treatment HIV-1 drug resistance

Of 3567 sequences from ART-naïve individuals, 550 (15.4%, 95% CI: 14.2–16.6) had at least one pre-treatment HIVDR mutation. NNRTI resistance mutations were most common (n=453, 12.7% [95% CI: 11.6–13.8), followed by NRTI (n=232, 6.5% [95% CI: 5.7–7.4]) and PIs (n=23, 0.9% [95% CI: 0.6–1.4). There were no INSTI resistance mutations observed over the analysis period, although the number of sequences analysed were also limited (n=106) (Figure 2a, and Table S1).

**Figure 2.**
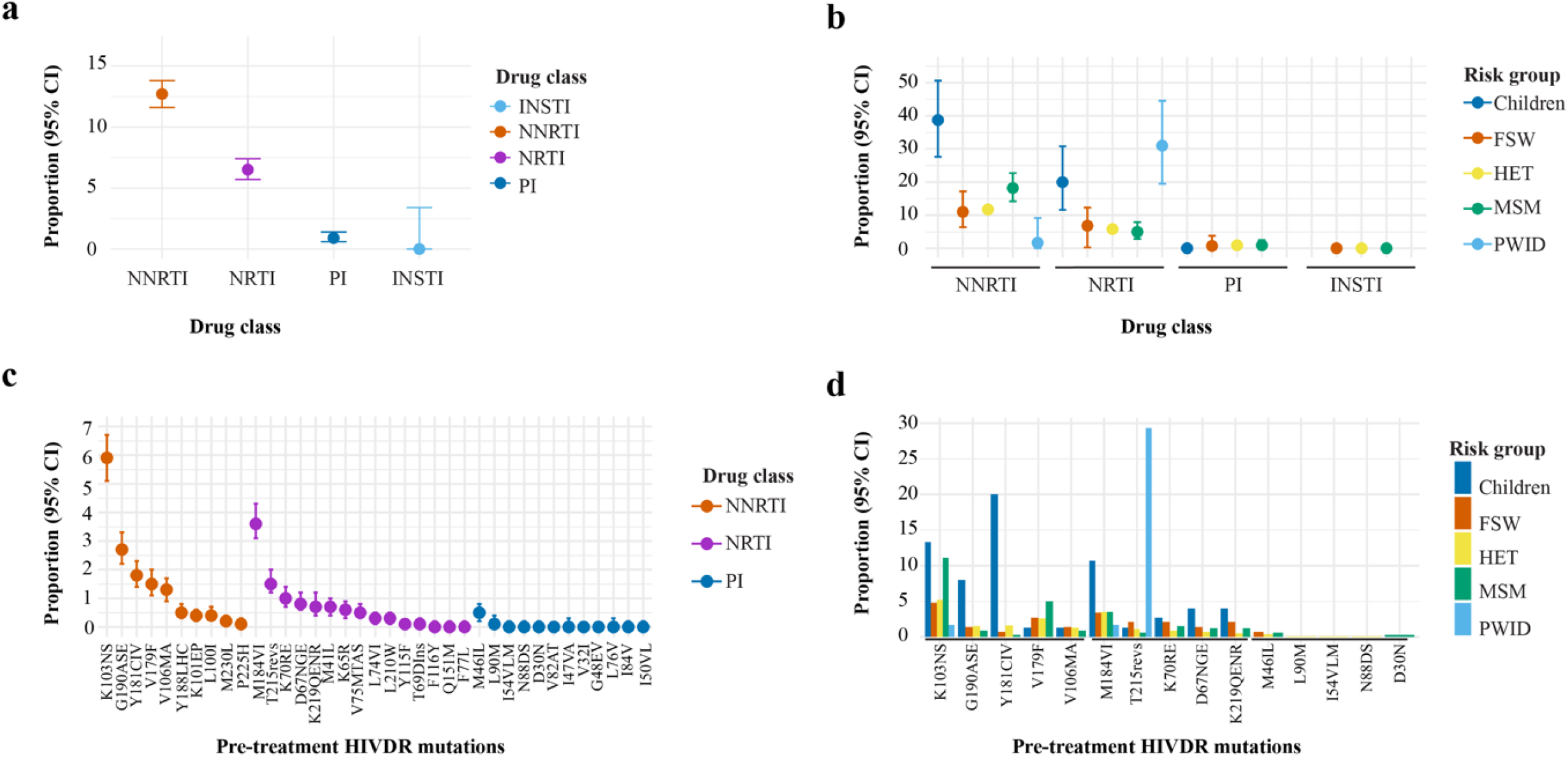
Distribution of pre-treatment HIV-1 drug resistance by (a) Proportion (95% CI) of ART-naive individuals with at least one HIV drug resistance mutations distributed by drug-class (for INSTI, CI is one-sided); (b) Distributed by drug class and risk group; (c) specific pre-treatment HIVDR mutations distributed by drug class; and (d) the five most prevalent pre-treatment HIVDR mutations per drug class distributed by risk groups, in Kenya (n=3567, 1986-2020).

Pre-treatment HIVDR mutations were most common amongst children (n=31, 41.3%), followed by PWID (n=18, 31.0%), MSM (n=68, 20.0%), FSW (n=22, 15.1%) and HET (n=411, 13.9%). Compared to other risk groups, children had higher pre-treatment NNRTI resistance mutations (n=29, 38.7%) while PWID had higher pre-treatment NRTI resistance mutations (n=18, 31.0%). Pre-treatment PI resistance mutations were <1.0% in HET, FSW, and MSM. Pre-treatment PI mutations were not observed in children and PWID although few sequences from these groups were analysed in this study (Figure 2b, and Table S1).

Overall, the most common pre-treatment NNRTI, NRTI and PI drug resistance mutations were the K103N/S (n=210, 5.9% of all sequences with any pre-treatment HIVDR), M184V (n=130, 3.6%) and M46I/L (n=12, 0.5%), respectively (Figure 2c, and Table S2). Whilst K103N/S was the most common NNRTI mutation amongst MSM, HET, FSW and PWID, the Y181C/I/V mutation was the most common amongst children. The T215revs were the most common NRTI mutations amongst PWID, whilst M184V/I was the most common NRTI mutation amongst children, FSW, HET and MSM (Figure 2d, and Table S2).

### Temporal trends in pre-treatment HIV-1 drug resistance

Overall, there was an increase in pre-treatment HIVDR from 6.9% (before 2005) to 24.2% (2016-2020, p□<□0.001). Pre-treatment NNRTI resistance increased from 3.8% (before 2005) to 22.9% (2016-2020, p<□0.001) while pre-treatment NRTI resistance increased from 4.2% (before 2005) to 8.1% (2016-2020, p=0.061). Pre-treatment PI resistance remained stable and below 2% throughout the calendar periods (p=0.298) (Figure 3, and Table S3).

**Figure 3.**
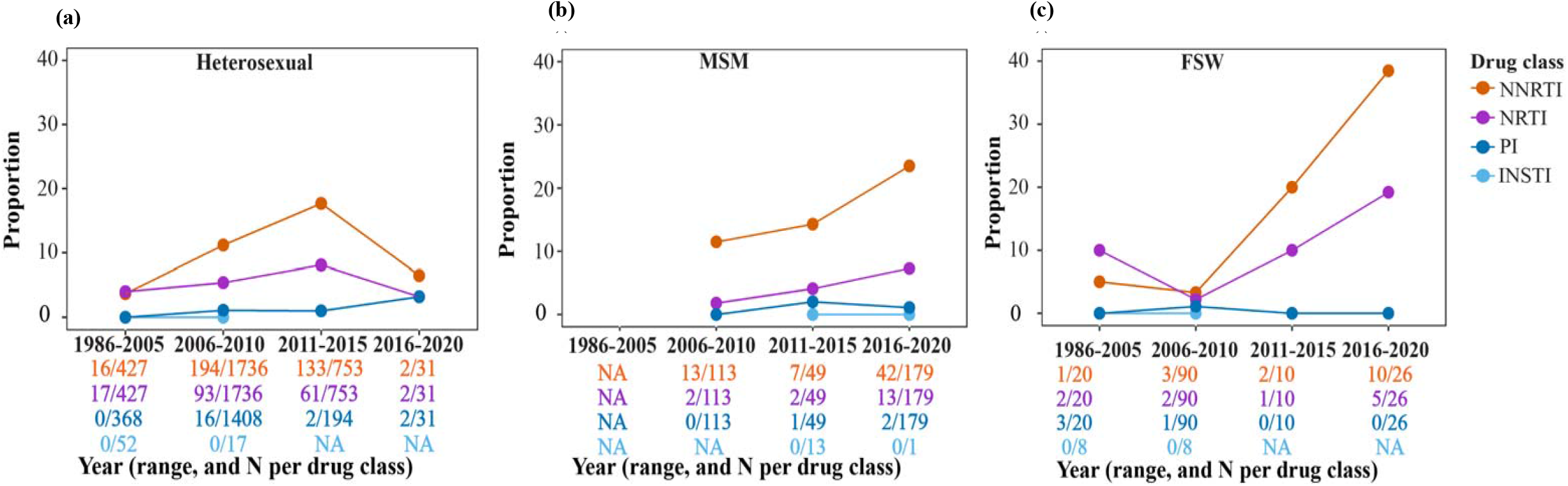
Temporal trends in pre-treatment HIV-1 drug resistance in different risk groups in Kenya (1986-2020). Overall trends and risk-group specific trends in pre-treatment HIV-1 drug resistance among (a) ART-naïve heterosexuals in the general population (HET), (b) men who have sex with men (MSM), and (c) female sex workers (FSW) in Kenya.

Amongst the HET population, overall pre-treatment HIVDR increased from 6.6% (before 2005) to 20.2% (2011-2015) – followed by a significant decline to 6.5% (2016-2020, p□<□0.001, LBL). Levels of pre-treatment NNRTI and NRTI resistance also increased from 3.7% and 4.0% (before 2005) to 17.7% and 8.1% (2011-2015), followed by a decline to 6.5% and 3.2% (2015-2020, p□<□0.001, and p=0.079, LBL respectively). Pre-treatment PI resistance remained low at 0.0% (before 2005), 1.0% (2011-2015) and 3.2% (2016-2020, p=0.839), though this was based on few sequences and hence overlapping binomial confidence intervals.

Among MSM there was an overall increase in NNRTI and NRTI resistance between 2006-2010 and 2015-2020, though this did not achieve statistical significance (p=0.399, and p=0.241, LBL respectively). Likewise, among FSW, NNRTI resistance also increased between 2006-2010 and 2015-2020 (p=0.001, LBL). Data on PWID and children fell within one calendar year grouping (2006-2010 and 2011-2015, respectively), and were therefore excluded from trend analysis (Table S3).

### Phylogenetic clustering of sequences with shared pre-treatment HIV-1 drug resistance mutations

Overall, 491 sequences with pre-treatment HIVDR mutations were analysed, of which 377 were subtype A1, subtype C (n=31), and subtype D (n=83). Cluster analysis revealed 32 clusters (size range 2-16) with the shared HIV-1 pre-treatment mutations K103N (n=14, 43.6% of all clusters), Y181C (n=6, 18.9%), M184V (n=5, 15.6%), G190A/S (n=3, 9.4%), and T215revs (n=4, 12.5%, Table S4).

The clusters were dyads (n=26, 84.5%), networks (n=4, 12.5%), and large clusters (n=1, 3.0%). Clusters were either risk-group exclusive i.e., HET only (n=24, 75.0%), MSM only (n=2, 6.3%), and PWID only (n=1, 3.1%), or mixed clusters including HET/MSM (n=2, 6.3%), HET/FSW (n=1, 3.1%), and HET/Children (n=2, 6.3%) (Table 3).

**Table 3.**
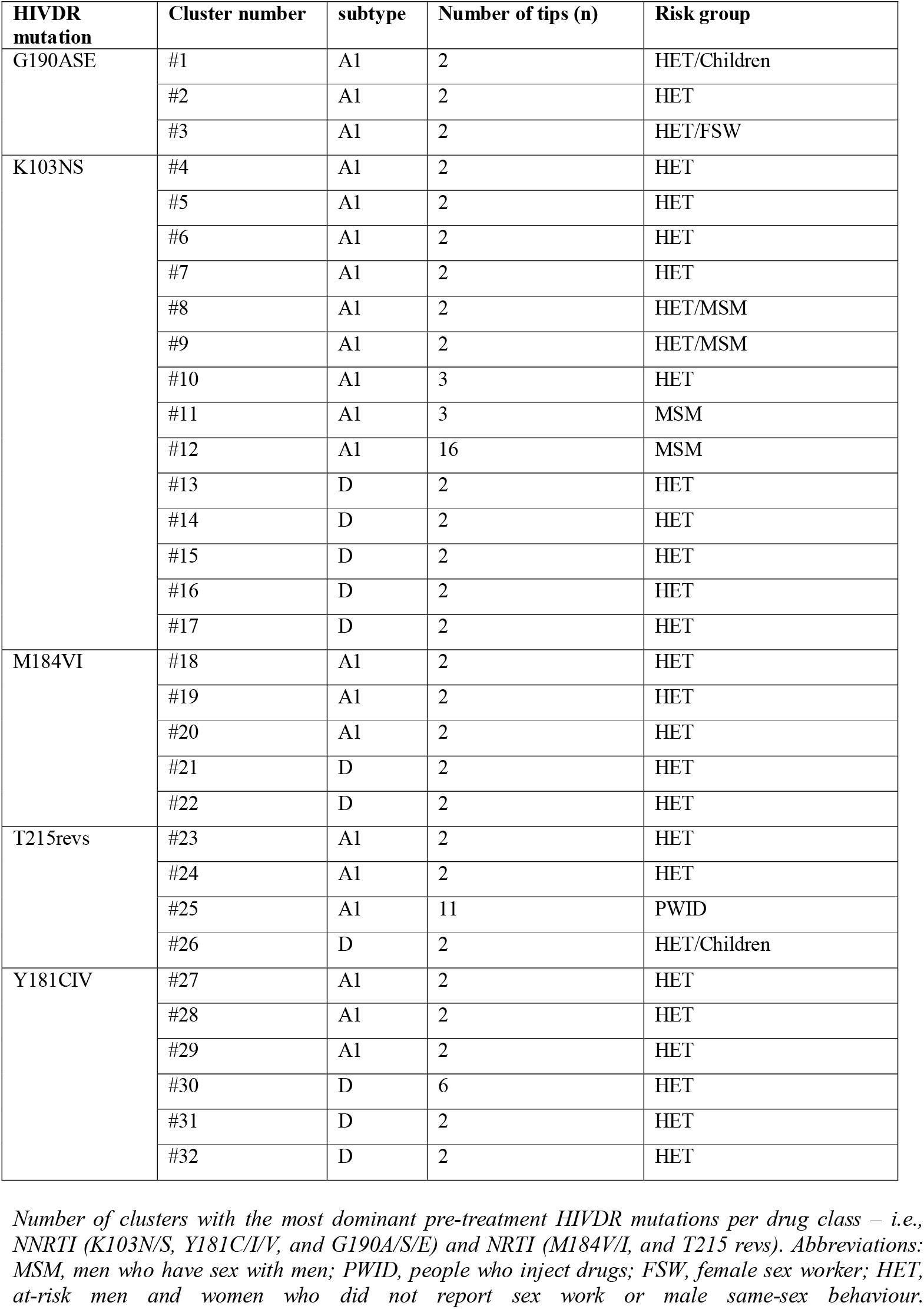
Characteristics of clusters (n=32) with shared pre-treatment HIVDR mutations and distributed into subtypes and risk group.

| HIVDR mutation | Cluster number | subtype | Number of tips (n) | Risk group |
| --- | --- | --- | --- | --- |
| G190ASE | #1 | A1 | 2 | HET/Children |
|  | #2 | A1 | 2 | HET |
|  | #3 | A1 | 2 | HET/FSW |
| K103NS | #4 | A1 | 2 | HET |
|  | #5 | A1 | 2 | HET |
|  | #6 | A1 | 2 | HET |
|  | #7 | A1 | 2 | HET |
|  | #8 | A1 | 2 | HET/MSM |
|  | #9 | A1 | 2 | HET/MSM |
|  | #10 | A1 | 3 | HET |
|  | #11 | A1 | 3 | MSM |
|  | #12 | A1 | 16 | MSM |
|  | #13 | D | 2 | HET |
|  | #14 | D | 2 | HET |
|  | #15 | D | 2 | HET |
|  | #16 | D | 2 | HET |
|  | #17 | D | 2 | HET |
| M184VI | #18 | A1 | 2 | HET |
|  | #19 | A1 | 2 | HET |
|  | #20 | A1 | 2 | HET |
|  | #21 | D | 2 | HET |
|  | #22 | D | 2 | HET |
| T215revs | #23 | A1 | 2 | HET |
|  | #24 | A1 | 2 | HET |
|  | #25 | A1 | 11 | PWID |
|  | #26 | D | 2 | HET/Children |
| Y181CIV | #27 | A1 | 2 | HET |
|  | #28 | A1 | 2 | HET |
|  | #29 | A1 | 2 | HET |
|  | #30 | D | 6 | HET |
|  | #31 | D | 2 | HET |
|  | #32 | D | 2 | HET |

*Number of clusters with the most dominant pre-treatment HIVDR mutations per drug class – i.e., NNRTI (K103N/S, Y181C/I/V, and G190A/S/E) and NRTI (M184V/I, and T215 revs). Abbreviations: MSM, men who have sex with men; PWID, people who inject drugs; FSW, female sex worker; HET, at-risk men and women who did not report sex work or male same-sex behaviour.*

Bayesian dating was performed for clusters with ≥3 sequences with shared pre-treatment HIVDR mutations – and included four subtype A1 clusters and one subtype D cluster (Table 4). The largest of these comprised 16 MSM having the K103N mutation. This cluster had an estimated tMRCA of 2005 (HPD, 2000 – 2008) where the most recent sample was collected in 2017, suggesting that this lineage had persisted over 12 years (growth rate = 0.1/year and R_0_=1.1) among ART naïve MSM in Kenya. Another long-standing pre-treatment HIVDR cluster involved 11 PWID having the T215rev mutation with an estimated tMRCA of 1999 (HPD, 1998 – 2001) where the most recent sample was collected in 2010 – indicating that this lineage had persisted (growth rate = 0.38/year, R_0_=1.38) over 11 years among ART-naïve PWID in Kenya. Overall, all except one pre-treatment HIVDR clusters had basic reproductive number, R_0_ >1, indicative of onward propagation of pre-treatment HIVDR mutations among untreated individuals in Kenya (Table 4).

**Table 4.** Characteristics of clusters having ≥3 sequences (n=5) with shared pre-treatment HIVDR mutations.

| Cluster | HIVDR mutation | Subtype | Tips (N) | TMRCa (95% HPD) | Sampling period | Growth rate | $R_0$<br>(where $D=1$) | Risk group |
| --- | --- | --- | --- | --- | --- | --- | --- | --- |
| #12.A1 | K103N | A1 | 16 | 2005 (2000 - 2008) | 2010-2017 | 0.10 | 1.10 | MSM |
| #30.D | Y181C | D | 6 | 2003 (1995 – 2006) | 2007-2013 | 0.00 | 1.00 | HET |
| #10.A1 | K103N | A1 | 3 | 2007 (2002 – 2010) | 2011-2015 | 0.94 | 1.94 | HET |
| #11.A1 | K103N | A1 | 3 | 2006 (2000 – 2006) | 2006-2007 | -0.06 | 0.94 | MSM |
| #25.A1 | T215revs | A1 | 11 | 1999 (1998 – 2001) | 2010 | 0.38 | 1.38 | PWID |

*Bayesian dating was restricted to 5 clusters having $\geq 3$ sequences and with the three most dominant pre-treatment HIVDR mutation per drug class – i.e., NNRTI (K103N/S, Y181C/I/V, and G190A/S/E) and NRTI pre-treatment HIVDR mutations (M184V/I, T215 revs, and K70R/E). Abbreviations: MSM (men who have sex with men); PWID (people who inject drugs); FSW (female sex workers); HET (at-risk men and women who did not report sex work or male same-sex behaviour); TMRCa (time to the most recent common ancestor); and HPD (higher posterior density interval).*

## DISCUSSION

We used HIV-1 *pol* sequence data from Kenya and spanning a period of more than three decades to assess the prevalence, temporal trends, and transmission of HIV-1 pre-treatment drug resistance among different risk groups in Kenya. Overall pre-treatment HIVDR prevalence was high (15.4%) – and these estimates were largely a reflection of the high levels of pre-treatment NNRTI drug resistance mutations. Notably, there was comparatively lower (<10%) pre-treatment NRTI drug resistance and no pre-treatment INSTI resistance in the study population (based on a limited number of HIV-1 *integrase* gene sequences), justifying the retention of two NRTIs – and a switch to a dolutegravir based-regimen for HIV-1 treatment in Kenya^45^.

We observed an increasing trend in pre-treatment HIVDR at a countrywide level, which was largely driven by NNRTI resistance, an observation that is consistent with previous sub-national data from Kenya and data from elsewhere in the global context^12-15,46^. NNRTIs have a low genetic barrier which permits outgrowth of resistant variants under sub-optimal drug pressure, and mutations may persist for long durations, facilitating their onward transmission^47,48^. Of interest, pre-treatment NNRTI and NRTI resistance among HET (by far the largest risk group in this study) increased between 2005 and 2015 but declined between 2015 and 2020. This coincides with the nationwide transition from NNRTI to INSTI-based ART regimens^45^ – and suggests that the reduction of the use of NNRTI-based regimens and the replacement with dolutegravir-based regimen may have abrogated emergence and circulation of pre-treatment NNRTI and NRTI resistance mutations. In contrast to trends among HET, pre-treatment HIVDR among FSW and MSM increased consistently through to 2015-2020. Remarkably, as of 2020, ART coverage was lower among key populations: 73% in FSW, 68% in PWID, and 63% in MSM, compared to 86% in the general HET population^49^. This may explain the higher resistance levels observed in key populations compared to HET, and requires further investigation. Enhanced monitoring of INSTI resistance using contemporary sequences from both HET and key populations would be prudent to inform future treatment strategies.

The high levels of K103N (in addition to Y181C, and G190A) mutations in this study likely reflect extensive selection of these mutations in persons receiving NVP and EFV – both of which comprised the main NNRTI options for first-line triple therapy regime historically in Kenya^50-53^. Also, we observed several HIV-1 clusters with shared mutations spanning more than 10 years, indicating onward propagation of HIVDR among treatment naïve individuals as new infections^54^. There was more frequent HIV-1 clustering of K103N strains. The largest cluster also comprised the K103N mutation, indicating extensive transmission of this pre-treatment mutation in Kenya, possibly dating back to the single-dose nevirapine era, as has been reported in other settings^55,56^. All except one of the clusters had a basic reproductive number of more than one, suggesting persistent NRTI and NNRTI HIVDR transmissions. Assessment of these trends over the next few years is needed to determine whether roll out of the more efficacious INSTI as a first line regimen will terminate circulation of NNRTI resistance mutations in Kenya. Also, all clusters were risk-group specific, consistent with previous reports that HIV-1 transmission in Kenya occurs mostly within risk groups, where HIV-1 mixing between risk groups is rare^21,22^.

The main strength of our study was the near-national level coverage and the ability to assess for temporal trends, dynamics, and transmission linkages spanning over three decades. However, the main limitation relates to the distribution of sequence data across time periods and by risk group, with some groups and time periods severely limited with very sparse sampling. Thus, temporal trends from drug-class estimates (more so for PI and INSTI mutations) and risk-group estimates (more so for children and PWID) should be interpreted with caution.

In conclusion, we demonstrated an increase in pre-treatment HIVDR over the last two decades, which justifies the switch to INSTI-based therapy in Kenya. We also report long-standing propagation of pre-treatment HIVDR mutations, but this might have been mitigated by the introduction of INSTI-based regimens. The switch to INSTI-based regimens may also have resulted in the abrogation of NRTI and NNRTI mutations, as observed in the general HET population. Our study further underscores the challenges with access to care and treatment for key populations as demonstrated with increasing pre-treatment HIVDR in MSM and FSW – despite the broad nationwide introduction of INSTI-based regimens. Taken together, our findings advocate for interventions for key populations towards equitable access to care and treatment, and continued vigilance for pre-treatment HIVDR surveillance to include INSTIs towards informing treatment options in Kenya.

## Data Availability

Nucleotide sequence data are available from GenBank under the accession numbers OM109695-OM110282

## Acknowledgments

We thank the staff affiliated with the MSM Health Research Consortium (MHRC) and the International AIDS Vaccine Initiative (IAVI) for supporting studies involving key populations in Kenya. This manuscript was submitted for publication with the permission from the Director of the Kenya Medical Research Institute (KEMRI).

## Author contributions

A.S.H., J.E and E.J.S. conceptualized, designed and provided funding for the study. E.J.S., S.M.G., J.K, L.M, F.C, M.M, K.M, G.M, M.M, O.A, L.G, A.D.S, and R.B provided samples from which new sequences used in the study were generated. G.N.M and Y.S performed lab work. G.M.N performed inferential analyses and produced figures and tables. M.K helped with producing figures and tables. D.A, and P.M helped with data analysis. G.N.M wrote the original draft manuscript and all the authors reviewed and edited the manuscript before submission.

