## Supplemental material for "TEMPORAL TRENDS AND TRANSMISSION DYNAMICS OF PRE-TREATMENT HIV-1 DRUG RESISTANCE WITHIN AND BETWEEN RISK GROUPS IN KENYA, 1986-2020"

^1^Lund University, Lund, Sweden, ^2^KEMRI/Wellcome Trust Research Programme, Kilifi, Kenya, ^3^Nyanza Reproductive Health Society, Kisumu, Kenya, ^4^University of Nairobi, Nairobi, Kenya, ^5^University of Manitoba, Winnipeg, Canada, ^6^Centre for the AIDS Programme of Research in South Africa (CAPRISA), South Africa, ^7^National Microbiology Laboratory at the JC Wilt Infectious Diseases Research Centre, Public Health Agency of Canada, Winnipeg, Canada, ^8^Pwani University, Kilifi, Kenya, ^9^KEMRI/Center for Global Health Research, Kisumu, Kenya, ^10^KAVI Institute of Clinical Research, University of Nairobi, Nairobi, Kenya, ^11^University of Washington, Seattle, USA, ^12^IAVI, New York, USA, ^13^University of California, San Francisco, USA, ^14^University of Illinois at Chicago, USA, ^15^University of Oxford, Oxford, United Kingdom.

**Files in this Data Supplement:**

Table S1. Frequency of drug class-specific pre-treatment HIV drug resistance mutations in different risk groups in Kenya (n=3567, 1986-2020).

Table S2. Frequency of drug class-specific pre-treatment HIVDR mutations by drug class and risk group among ART-naïve individuals in Kenya (n=3567, 1986-2020).

Table S3. Temporal trends in HIV-1 drug resistance among ART naïve individuals with different risk groups in Kenya (1986-2020).

### Table S1. Frequency of drug class-specific pre-treatment HIV drug resistance mutations in different risk groups in Kenya, including the frequencies, (proportions and 95% confidence intervals [95% CI]) (n=3567, 1986-2020).

| **Drug class** | **Overall** | **HET** | **MSM** | **FSW** | **PWID** | **Children** |
| --- | --- | --- | --- | --- | --- | --- |
| Any HIVDR | 550/3567  (15.4 [14.2-16.6]) | 411/2947  (13.9 [12.7-15.2]) | 68/341  (20.0 [15.8-24.6]) | 22/146  (15.1 [9.7-21.9]) | 18/58  (31.0 [19.5-44.5]) | 31/75  (41.3 [30.1-53.3]) |
| NNRTI | 453/3567  (12.7 [11.6-13.8]) | 345/2947  (11.7 [10.6-12.9]) | 62/341  (18.2 [14.2-22.7]) | 16/146  (11.0 [6.4-17.2]) | 1/58  (1.7 [0-9.2]) | 29/75  (38.7 [27.6-50.6]) |
| NRTI | 232/3567  (6.5 [5.7-7.4]) | 172/2947  (5.8 [5.0-6.7]) | 17/341  (5.0 [2.9-7.9]) | 10/146  (6.8 [0.3-12.3]) | 18/58  (31.0 [19.5-44.5]) | 15/75  (20.0 [11.6-30.8]) |
| PI | 23/2491  (0.9 [0.6-1.4]) | 19/2001  (0.9 [0.6-1.5]) | 3/341  (0.9 [0.1-2.5]) | 1/146  (0.7 [0-3.8]) | (N/A) | 0/3  (0.0 [0-70.8]) |
| INSTI | 0/106  (0.0 [0.0-3.4^*^]) | 0/69  (0.0 [0.0-0.5^*^]) | 0/21  (0.0 [0.0-16.1^*^]) | 0/16  (0.0 [0.0-20.6^*^]) | (N/A) | (N/A) |

*Abbreviations: ART, antiretroviral therapy; HET, presumed heterosexual i.e., men and women not reporting sex work or male same-sex behaviour; MSM, men who have sex with men; PWID, people who inject drugs; FSW, female sex worker; NRTI, nucleoside reverse transcriptase inhibitors; NNRTI, non-nucleoside reverse transcriptase inhibitors; PI, protease inhibitors; INSTI, integrase strand transfer inhibitors. *One-sided, 97.5% confidence interval.*

### Table S2. Frequency and proportion of drug class-specific pre-treatment HIVDR mutations among sequences from treatment naïve with any pre-treatment HIVDR mutation by drug class and risk group among ART-naïve individuals in Kenya (n=3567, 1986-2020).

| **Drug class** | **HIVDR mutation** | **Number and proportion** |
| --- | --- | --- |
| NNRTI | K103NS | 210 (5.9%) |
|  | G190ASE | 98 (2.7%) |
|  | Y181CIV | 65 (1.8%) |
|  | V179F | 55 (1.5%) |
|  | V106MA | 45 (1.3%) |
|  | Y188LHC | 17 (0.5%) |
|  | K101EP | 16 (0.4%) |
|  | L100I | 13 (0.4%) |
|  | M230L | 8 (0.2%) |
|  | P225H | 5 (0.1%) |
| NRTI | M184VI | 130 (3.6%) |
|  | T215revs | 56 (1.6%) |
|  | K70RE | 36 (1.0%) |
|  | D67NGE | 30 (0.8%) |
|  | K219QENR | 24 (0.7%) |
|  | M41L | 20 (0.6%) |
|  | K65R | 18 (0.5%) |
|  | V75MTAS | 10 (0.3%) |
|  | L74VI | 9 (0.3%) |
|  | L210W | 5 (0.1%) |
|  | Y115F | 3 (0.1%) |
|  | T69DIns | 3 (0.1%) |
|  | F116Y | 2 (0.1%) |
|  | Q151M | 2 (0.1%) |
|  | F77L | 1 (0.0%) |
| PI | M46IL | 12 (0.5%) |
|  | L90M | 3 (0.1%) |
|  | I54VL | 2 (0.1%) |
|  | N88DS | 2 (0.1%) |
|  | D30N | 2 (0.1%) |
|  | V82A | 1 (0.0%) |
|  | I47VA | 1 (0.0%) |
|  | V32I | 1 (0.0%) |
|  | G48E | 1 (0.0%) |
| INSTI | None | None |

*Abbreviations: ART, anti-retroviral therapy; NRTI, nucleoside reverse transcriptase inhibitors; NNRTI, non-nucleoside reverse transcriptase inhibitors; PI, protease inhibitors; INSTI, integrase strand transfer inhibitors.*

### Table S3. Temporal trends in HIV-1 drug resistance among ART naïve individuals with different risk groups, including the frequencies, (proportions and 95% confidence intervals [95% CI]) in Kenya (1986-2020).

| **Risk group** | **Years** | **Any HIVDR** | **NNRTI** | **NRTI** | **PI** | **INSTI** |
| --- | --- | --- | --- | --- | --- | --- |
| Overall | Before 2005 | 31/451  (6.9 [4.7-9.6]) | 17/451  (3.8 [2.2-6]) | 19/451  (4.2 [2.6-6.5]) | 0/391  (0.0 [0.0-0.9^*^]) | 0/60  (0.0 [0.0-6.0^*^]) |
|  | 2006-2010 | 268/1997  (13.4 [12-15]) | 211/1997  (10.6 [9.3-12]) | 115/1997  (5.8 [4.8-6.9]) | 17/1611  (1.1 [0.6-1.6]) | 0/42  (0.0 [0.0-8.4^*^]) |
|  | 2011-2015 | 194/883  (22.0 [19.3-24.8]) | 171/883  (19.4 [16.8-22.1]) | 79/883  (8.9 [7.1-11]) | 3/253  (1.1 [0.2-3.4]) | 0/4  (0.0 [0.0-60.2^*^]) |
|  | 2016-2020 | 57/236  (24.2 [18.8-30.1]) | 54/236  (22.9 [17.7-28.8]) | 19/236  (8.1 [4.9-12.3]) | 3/236  (1.3 [0.3-3.7]) | (N/A) |
| HET | Before 2005 | 28/427  (6.6 [4.4-9.3]) | 16/427  (3.7 [2.2-6]) | 17/427  (4.0 [2.3-6.3]) | 0/368  (0.0 [0.0-1.0]) | 0/52  (0.0 [0.0-6.8^*^]) |
|  | 2006-2010 | 229/1736  (13.2 [11.6-14.9]) | 194/1736  (11.2 [9.7-12.8]) | 93/1736  (5.4 [4.3-6.5]) | 16/1408  (1.1 [0.7-1.8]) | 0/17  (0.0 [0.0-19.5^*^]) |
|  | 2011-2015 | 152/753  (20.2 [17.4-23.2]) | 133/753  (17.7 [15-20.6]) | 61/753  (8.1 [6.3-10.3]) | 2/194  (1.0 [0.1-3.7]) | (N/A) |
|  | 2016-2020 | 2/31  (6.5 [0.8-21.4]) | 2/31  (6.5 [0.8-21.4]) | 1/31  (3.2 [0.1-16.7]) | 1/31  (3.2 [0.1-16.7]) | (N/A) |
| MSM | Before 2005 | (N/A) | (N/A) | (N/A) | (N/A) | (N/A) |
|  | 2006-2010 | 15/113  (13.3 [7.6-20.9]) | 13/113  (11.5 [6.3-18.9]) | 2/113  (1.8 [0.2-6.2]) | 0/113  (0.0 [0.0-3.2^*^]) | (N/A) |
|  | 2011-2015 | 8/49  (16.3 [7.3-29.7]) | 7/49  (14.3 [5.9-27.2]) | 2/49  (4.1 [0.5-14]) | 1/49  (2 [0.1-10.9]) | 0/13  (0.0 [0.0-24.7^*^]) |
|  | 2016-2020 | 45/179  (25.1 [19-32.2]) | 42/179  (23.5 [17.5-30.4]) | 13/179  (7.3 [3.9-12.1]) | 2/179  (1.1 [0.1-4.0]) | 0/1  (0.0 [0.0-97.5^*^]) |
| PWID | Before 2005 | (N/A) | (N/A) | (N/A) | (N/A) | (N/A) |
|  | 2006-2010 | 18/58  (31.0 [19.5-44.5]) | 1/58  (1.7 [0.0-9.2]) | 18/58  (31 [19.5-44.5]) | (N/A) | 0/58  (0.0 [0.0-6.2^*^]) |
|  | 2011-2015 | (N/A) | (N/A) | (N/A) | (N/A) | (N/A) |
|  | 2016-2020 | (N/A) | (N/A) | (N/A) | (N/A) | (N/A) |
| FSW | Before 2005 | 3/20  (15.0 [3.2-37.9]) | 1/20  (5.0 [0.1-24.9]) | 2/20  (10.0 [1.2-31.7]) | 3/20  (15.0 [3.2-37.9]) | 0/8  (0.0 [0.0-36.9]) |
|  | 2006-2010 | 6/90  (6.7 [2.5-13.9]) | 3/90  (3.3 [0.7-9.4]) | 2/90  (2.2 [0.3-7.8]) | 1/90  (1.1 [0.0-6.0]) | 0/8  (0.0 [0.0-36.9]) |
|  | 2011-2015 | 3/10  (30.0 [6.7-65.2]) | 2/10  (20 [2.5-55.6]) | 1/10  (10.0 [0.3-44.5]) | 0/10  (0.0 [0.0-30.8^*^]) | (N/A) |
|  | 2016-2020 | 10/26  (38.5 [20.2-59.4]) | 10/26  (38.5 [20.2-59.4]) | 5/26  19.2 (6.6-39.4) | 0/26  (0.0 [0.0-13.2^*^]) | (N/A) |
| Children | Before 2005 | 0/4  (0.0 [0.0-60.2^*^]) | 0/4  (0.0 [0.0-60.2^*^]) | 0/4  (0.0 [0.0-60.2^*^]) | 0/3  (0.0 [0.0-70.6^*^]) | (N/A) |
|  | 2006-2010 | (N/A) | (N/A) | (N/A) | (N/A) | (N/A) |
|  | 2011-2015 | 31/71  (43.6 [31.9-56.0]) | 29/71  (40.8 [29.3-53.2]) | 15/71  (21.1 [12.3-32.4]) | (N/A) | (N/A) |
|  | 2016-2020 | (N/A) | (N/A) | (N/A) | (N/A) | (N/A) |

*Abbreviations: MSM, men who have sex with men; PWID, people who inject drugs; FSW, female sex worker; HET, at-risk men and women who did not report sex work or male same-sex behaviour; NRTI, nucleoside reverse transcriptase inhibitors; NNRTI, non-nucleoside reverse transcriptase inhibitors; PI, protease inhibitors; INSTI, integrase strand transfer inhibitors.*

** One-sided, 97.5% confidence interval.*
